# Cognitive and Behavioral Functioning in Female Former Soccer Players: Results from the Head Impact and Trauma Surveillance Study (HITSS)

**DOI:** 10.64898/2026.02.25.26347083

**Authors:** Shania C. Mulayi, Anna Aaronson, Kelsey J. Goostrey, Fatima Tuz-Zahra, Yorghos Tripodis, William S. Cole-French, Matthew Roebuck, Greta Schneider, Brittany N. Pine, Joseph N. Palmisano, Brett M. Martin, Kenton H. Zavitz, Douglas I. Katz, Christopher J. Nowinski, Ann C. McKee, Thor D. Stein, R. Scott Mackin, Michael D. McClean, Jennifer Weuve, Jesse Mez, Michael W. Weiner, Rachel L. Nosheny, Michael L. Alosco, Robert A. Stern

## Abstract

Repetitive head impacts (RHI) from contact and collision sports have been associated with later-life cognitive and neurobehavioral impairments, as well as neurodegenerative conditions such as chronic traumatic encephalopathy (CTE). RHI-associated clinical sequelae among female former soccer players, specifically, are not well understood. This cross-sectional study aimed to examine the relationship of RHI exposure proxies (e.g., total years of soccer play, concussion history, highest level of play, and estimated cumulative heading frequency) with clinical measures (e.g., subjective cognitive complaints, objective cognitive performance, behavioral dysregulation, and depressive symptoms) among 2,732 women, aged 40 years or above, enrolled in the Head Impact and Trauma Surveillance Study (HITSS), all of whom formerly played organized soccer. HITSS participants completed an online battery that elicited self-reported cognitive and behavioral complaints and depressive symptoms, and that assessed cognitive performing via computerized tests. Multivariable linear regression models estimated associations between soccer-related RHI proxies and outcome measures, adjusting for age and education. Among the former soccer players, longer duration of soccer play, higher level of play, greater estimated cumulative heading frequency, and concussion history were significantly associated with worse self-reported cognitive functioning, greater behavioral dysregulation, and elevated depressive symptom severity. Apart from a single association between concussion history and PAL FAMS performance, we found no other associations between RHI proxies and objective cognitive test performance. Among middle-aged women who played organized soccer, cumulative RHI exposure was associated with small but statistically significant effects for measures of subjective cognitive complaints, behavioral functioning, and depressive symptoms. Continued monitoring of this large cohort of female former soccer players will improve understanding of long-term consequences of soccer play.

## INTRODUCTION

Exposure to repetitive head impacts (RHI) from contact and collision sports has been linked to later-life cognitive and behavioral impairments,^1–4^ as well as neurodegenerative diseases, including chronic traumatic encephalopathy (CTE).^5,6^ Most of this research has focused on men who have played American football. A growing body of work has been investigating neuropathological, cognitive, and neuropsychiatric sequelae of soccer play. CTE has been pathologically confirmed in former soccer players,^7–9^ and a 2023 study found that elite male soccer players had a significantly elevated risk of neurodegenerative disease compared to non-RHI-exposed controls.^10^ In vivo studies, primarily of men, have also demonstrated associations between soccer play and cortical thinning, white matter disruptions,^11,12^ altered neurochemistry,^13^ and lower performance on neuropsychological tests, including measures of psychomotor speed and verbal memory.^14^

Heading the soccer ball has been associated with cognitive deficits and neurophysiological changes among current and former soccer players, although findings have been mixed. Short term exposure (e.g., immediately following a heading session, or heading in subsequent weeks or seasons) has been tied to poorer psychomotor-speed performance and slower reaction times on executive function tasks,^15^ worse performance on a measure of working memory,^16^ and elevated blood-based biomarkers of neural damage (p-Tau217 and S100B).^17^ However, other studies found no acute deficits on an anticipation timing test,^18^ and no differences in concentrations of biomarkers of neuronal injury in individuals who headed the ball versus those who did not.^19^ Long-term heading exposure has been linked to poorer verbal learning and memory,^20^ as well as white matter microstructural alterations,^21^ including lower fractional anisotropy in temporo-occipital white matter areas.^22^ Additional studies have observed greater neurodegenerative disease risk among male former professional soccer players with longer career duration and those in non-goalkeeper positions,^23^ and former professional players also have higher neurodegenerative disease mortality compared to matched controls.^24^ Quantifying risk in soccer play, and characterizing clinical symptoms associated with cumulative RHI exposure, however, remains challenging.

Most existing literature on the short and long-term consequences of soccer participation has focused on male soccer players, leaving female players comparatively under-included. Across all levels of play, female athletes have been shown to be at greater risk of sustaining head injuries.^25,26^ In soccer specifically, studies have found that female players experience worse post-concussion outcomes, including more symptoms and poorer visual and verbal memory,^27^ as well as more extensive white matter microstructural alterations in association with heading the ball.^28^ These differential responses may stem in part to sex-based anatomical differences, such as neck strength,^29^ as well as hormonal influences. For instance, fluctuations in estrogen and progesterone may modulate neuroinflammatory responses and cerebral metabolism post-head injury.^30^

When comparing female soccer players to non-contact sport athletes, some studies have reported that soccer players perform significantly worse on measures of verbal memory, psychomotor speed, and verbal fluency.^31,32^ However, other studies report no adverse cognitive effects when compared to controls.^33^ For example, one study found that female soccer players exhibited normal age-related cognitive trajectories across multiple domains, including memory, psychomotor speed, reaction time, complex attention and cognitive flexibility, and in some cases, performed better than normative samples.^34^ With millions of female soccer players in the United States alone, these mixed findings underscore the need for greater research focused specifically on cognitive and behavioral consequences of soccer participation among women.

This study examined associations between self-reported proxies for RHI exposure (e.g., duration of soccer participation, highest level of soccer play, estimated cumulative heading frequency, and concussion history) and objective cognitive performance, subjective cognitive complaints, self-reported behavioral dysregulation, and depressive symptoms among a large cohort of women who played organized soccer, age 40 years and older, whose highest level of play spanned youth, high school, collegiate, semi-professional, and professional play. We hypothesized that greater RHI exposure from soccer would be associated with poorer cognitive test performance, increased subjective cognitive decline, greater behavioral dysregulation, and elevated depressive symptoms.

## METHODS

### 2.1 Study Design

The sample included 2,732 women enrolled in the Head Impact and Trauma Surveillance Study (HITSS) who reported that they had previously played organized soccer. HITSS is a longitudinal, observational study examining cognitive and behavioral functioning in former soccer and American-style football players aged 40 years or above in the United States. HITSS is an extension of the University of California, San Francisco (UCSF) Brain Health Registry (BHR), an online Alzheimer’s disease and related dementias research registry for the recruitment and longitudinal monitoring of >105,000 adults.^35,36^ HITSS includes a remote online battery composed of questionnaires eliciting self-reported data on demographics, athletic and medical history, and cognitive and behavioral complaints; the battery also includes three computerized cognitive tests. The battery takes approximately 90 minutes to complete, and, every twelve months, HITSS invites participants to complete subsequent waves of assessments via automated email messages. Participants receive no monetary compensation for task completion but may enter a $500 gift card drawing. Recruitment of former soccer players was conducted via four channels: digital advertising (e.g., via Facebook and Instagram), organic media, such as press releases issued to large-scale media publications, in-person recruitment, and organizational partnerships. All participants provided written, electronic informed consent, and the study is approved by the Boston University Medical Campus Institutional Review Board. HITSS began enrolling participants in March 2022.

### 2.2 Sample

The study consisted of a total of 2,732 female former soccer players spanning multiple levels of play: professional/semi-professional, college, high school, and youth. Participants were included if they self-identified as female, reported soccer as their primary sport (e.g., sport with the greatest duration of play), indicated that they no longer participated in soccer, and completed at least one of our seven outcomes of interest in their baseline evaluation before April 9, 2025. Participants were excluded if they played the following contact and collision sports: American football, hockey, boxing, futsal, rugby, wrestling, mixed martial arts, bull riding, equestrian, auto racing, horse jumping, or roller hockey. Completion rates by highest level of play were determined for each of the outcome measures. (**Table 1).**

**Table 1:**
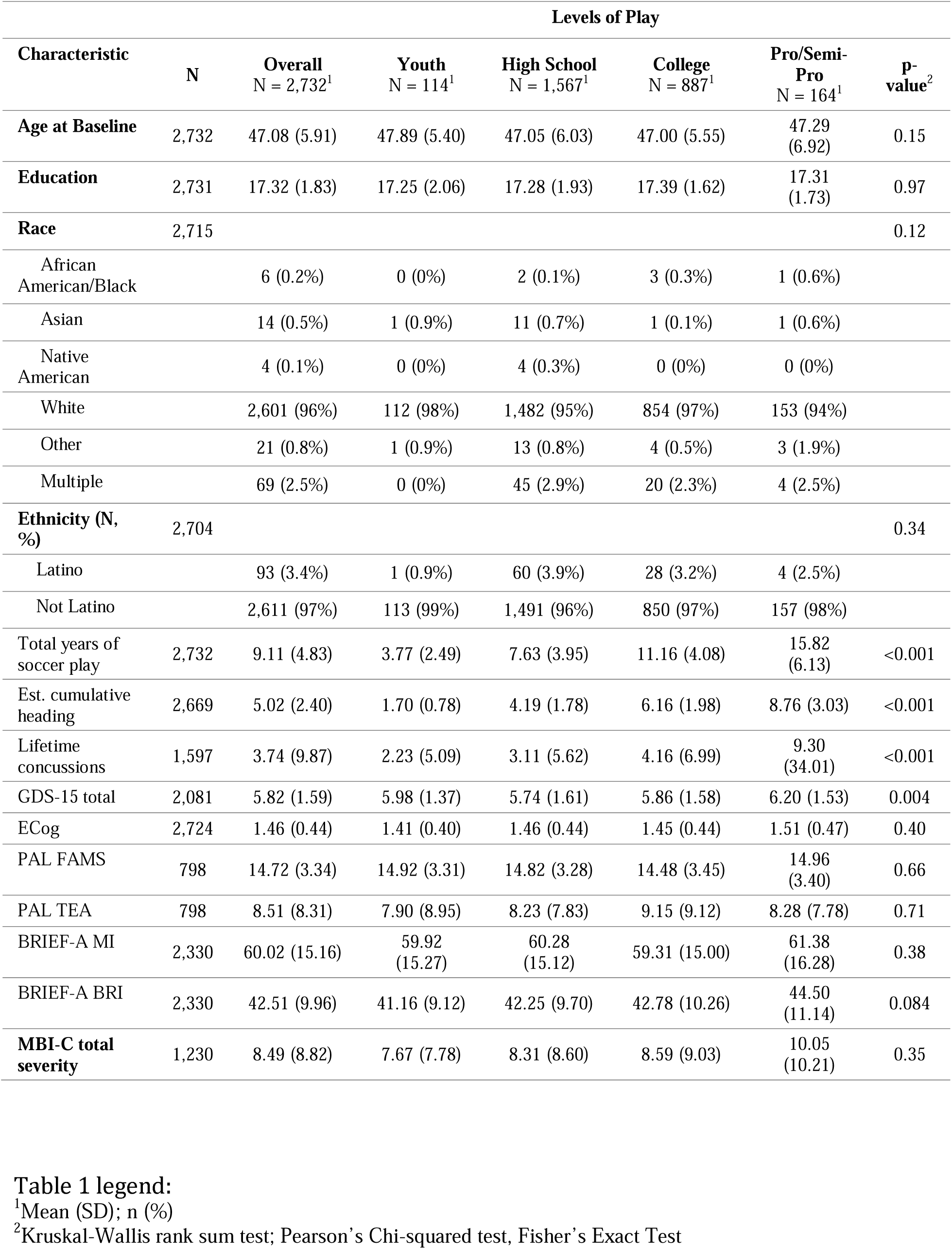
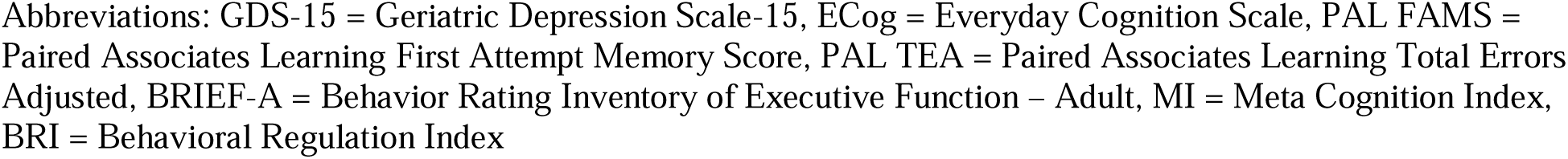
Soccer Player Characteristics.

### 2.3 Measures

#### 2.3.1 Measures of RHI Exposure

##### Boston University Repetitive Head Impact Exposure Assessment (RHIEA)

Investigators at Boston University developed the RHIEA to evaluate sports participation and other sources of RHI such as military service.^37–39^ Participants are asked: “Have you ever participated in organized sports, which includes membership on a team with scheduled practices and games (excluding pick-up or neighborhood games)?” If the participant answers yes, they are asked additional questions regarding their participation in sports. If no, participants were directed to the next assessment; we excluded such participants from these analyses.

In this study, four metrics of RHI exposure were examined using the RHIEA: 1) Estimated cumulative heading exposure, derived by assigning numeric values to responses to the following question: “On average, compared to others who played the same position as you, how often did you head the ball?” Responses included: Less Frequently = 1, As Frequently = 2, and More Frequently = 3. These values were then summed across each participant’s total levels of play. This derivation included five levels of play (youth, high school, college, semi-professional and professional, and therefore ranged from 0-15); 2) Total years of play, which is defined as the total number of years the participant played organized soccer; 3) Highest level of play (i.e., youth [YTH], high school [HS], college [COL], or professional [PRO], with professional defined as having played on a national team, or in a professional or semi-professional league. 4) Concussion History determined from participant responses to the following question: “As best as you can remember, how many total concussions did you have during your life?” To ensure accurate and standardized reporting, participants were provided with the clear and accessible definition of concussion: “a concussion has occurred anytime you have had a blow to the head that caused you to have symptoms for any amount of time. These include: blurred or double vision, seeing stars, sensitivity to light or noise, headache, dizziness or balance problems, nausea, vomiting, trouble sleeping, fatigue, confusion, difficulty remembering, difficulty concentrating, or loss of consciousness. Whenever anyone gets a “ding” or their “bell rung,” that too is a concussion.” Responses were recorded as a continuous numeric variable of self-reported lifetime concussions.

#### 2.3.2 Subjective Cognitive Complaints

##### Behavior Rating Inventory of Executive Function- Adult version (BRIEF-A) Metacognition Index (MI)

The BRIEF-A is a widely used measure of executive functioning and self-regulation in daily life, based on 75 self-report items.^40^ Participants rate each item on a 3-point Likert scale (1 = never, 2 = sometimes, 3 = often). There are nine clinical subscales, which are empirically validated and divided into two indices: Metacognition Index (MI) and Behavioral Regulation Index (BRI). The MI encompasses the Initiate, Working Memory, Plan/Organize, Organization of Materials, and Monitor clinical subscales. Standardized age- and education-based T-scores for each clinical scale and the MI were used in this study. 2,312 participants (142 PRO, 740 COL, 1330 HS, 101YTH) completed this task.

##### Everyday Cognition Scale (ECog)

The ECog is a 39-question assessment designed to evaluate a participant’s perception of changes in their ability to perform daily tasks compared to ten years prior across six cognitive domains: memory, language, visuospatial abilities, planning, organization, and divided attention.^41^ Participants rate each item on a 4-point Likert scale ranging from “better or no change” to “consistently much worse.” The ECog mean score for each participant was calculated by averaging their responses across all questions, with higher scores indicating greater self-reported cognitive decline. 2,725 participants (164 PRO, 884 COL, 1563 HS, and 114 YTH) completed this task.

#### 2.3.3 Objective cognitive performance (CANTAB)

##### Paired Associates Learning (PAL)

The PAL is part of the Cambridge Neuropsychological Test Automated Battery (CANTAB) evaluates episodic memory and visual learning.^42–44^ The task involves pairing different images and testing the participant’s ability to learn and recall the locations of various patterns. Over time, the difficulty increases, as the number of patterns increases. The total number of errors and the first attempt memory score (FAMS), which measures the initial memory performance, are recorded to assess short-term memory and immediate recall^18^. An online version of the PAL was used and 797 participants (46 PRO, 251 COL, 461 HS, and 39 YTH) completed this task.

##### One Touch Stockings (OTS)

The OTS^45^ is another CANTAB test, and used to assess executive functioning, specifically, spatial planning and problem-solving ability. During the test, participants view two arrangements of colored balls and must determine the minimum number of moves needed to match one to the other. Instead of manipulating the objects, they respond with a single touch, selecting how many moves would be required. The test measures planning ability, working memory, and decision-making under increasing levels of difficulty. This analysis examined OTS Problems Solved on First Choice (OTSPSFC), the total number of assessed trials in which the participant selected the correct answer on their first attempt. 596 participants (24 PRO, 185 COL, 358 HS, 29 YTH) completed this task.

##### Spatial Working Memory (SWM)

The CANTAB SWM^45^ test evaluates an individual’s ability to retain and manipulate spatial information, an aspect of executive function. Participants search for hidden tokens inside boxes on the screen, while avoiding boxes already checked, requiring the use of spatial memory and strategy. As the task becomes more complex, it increasingly challenges the participant’s working memory capacity. For this study, we included two SWM scores: (1) SWM Strategy (SWMS), the number of times a participant began a new search pattern from the same box they previously started with. If they always begin a search from the same starting point, it can be inferred that the participant is using a pre-planned strategy to find tokens. (2) SWM Between Errors (SWMBE): The number of times the participant incorrectly revisited a box in which a token was previously found. This metric is calculated across all four, six and eight token trials. 748 participants (39 PRO, 235 COL, 436 HS, 38 YTH) completed this task.

#### 2.3.4: Behavioral and Depressive Symptoms

##### BRIEF-A Behavioral Regulation Index (BRI)

The BRIEF-A Behavioral Regulation Index (BRI)^46^ is derived from 30 self-report items assessing an individual’s ability to monitor their behavior and regulate impulses. This index includes the Inhibit, Shift, Emotional Control, and Self-monitor subscales. The scores for each clinical scale and index are summed, and for this study, the raw scores for each subscale and index were converted into T-scores. 2,312 participants (142 PRO, 740 COL, 1330 HS, 101YTH) completed this task.

##### Geriatric Depression Scale (GDS-15)

The GDS-15 is a self-report screening tool used to identify symptoms of depression in older adults.^47^ This measure consists of 15 yes-or-no questions evaluating the emotional and cognitive aspects of depression. The number of “yes” responses are summed, with total scores ranging from 0 to 15. Normal scores range from 0 to 4 while scores ranging from 5 to 8 suggest mild depression, 9 to 11 indicates moderate depression, and 12 to 15 suggests severe depression. The GDS-15 has demonstrated good sensitivity and specificity among adults aged 18 and older.^48^ 2072 participants (127 PRO, 680 COL, 1174 HS, and 91 YTH) completed this task.

##### Mild Behavioral Impairment Checklist (MBI-C)

The MBI-C is a 34-item survey used to identify persistent behavioral and psychological change in older adults prior to cognitive decline and dementia.^49^ The checklist assesses five key domains: 1) Decreased motivation; 2) Emotional dysregulation; 3) Impulse dyscontrol; 4) Social inappropriateness; and 5) Abnormal perception or thought content. For each item, “yes” or “no” is used to indicate the presence of a symptom. If present, the symptom is rated based on severity using a 3-point Likert scale (1=mild, 2= moderate, 3=severe). A total severity score is calculated by taking the sum of the severity for all endorsed symptoms. Higher scores indicate a greater risk for mild cognitive impairment with a cut off of 8.5.^50^ 1916 participants (113PRO, 612 COL, 1113 HS, and 78 YTH) completed this task.

### 2.4 Statistical Analysis

All statistical analyses were performed using R version 4.5.1 and SPSS version 29.0.2.0. We fit multivariable-adjusted linear regression models to estimate associations of total years of soccer play and cumulative heading with each outcome measure. All estimates were adjusted for years of education. Additionally, all models using raw scores (when T-scores were unavailable) were adjusted for age in years. Analyses of covariance (ANCOVA) models estimated associations between highest level of soccer play (YTH, HS, COL, PRO), with each outcome measure. Omnibus tests were employed to ascertain the overall effect of highest level of soccer play on each outcome. Post hoc Tukey-adjusted pairwise comparisons compared estimated marginal means across each level of play.

P-values from the linear regression and ANCOVA models were adjusted by applying the Benjamini-Hochberg false discovery rate (FDR) method independently for each of the RHI proxies (level of play, years of play, estimated cumulative heading). The total number of hypotheses was set to two for the subjective cognitive domain (BRIEF-A MI, ECog) three for the mood/neurobehavioral domain (GDS, MBI-C, BRIEF-A BRI), and five for the objective cognitive (CANTAB) metrics (PALFAMS, PALTEA, SWMS, SWMBE, OTSPFC).

#### Sensitivity Analyses

All multivariable linear regression and ANCOVA models were performed excluding professional players (N=164). Additional models were performed to examine associations of each RHI exposure metric with each BRIEF-A subscale (Inhibit, Shift, Emotional Control, Self-Monitor, Initiate, Working Memory, Plan/Organize, Task Monitor, and Organization of Materials).

## RESULTS

### 3.1 Sample

The soccer cohort (N= 2,732) had a mean age of 47.08 years (SD=5.91), and predominantly identified as white (2,601, 96%), and non-Latino (N=2,611, 97%). **Table 1**. includes detailed sample characteristics. Sample derivation is described in **Figure 1**. There were only one significant association between the RHI proxies and performance on any of the CANTAB objective cognitive tests; concussion history and PAL FAMS. The following sections detail the findings on the self-report measures of subjective cognitive decline, depressive symptoms, and behavioral dysregulation.

**Figure 1:**
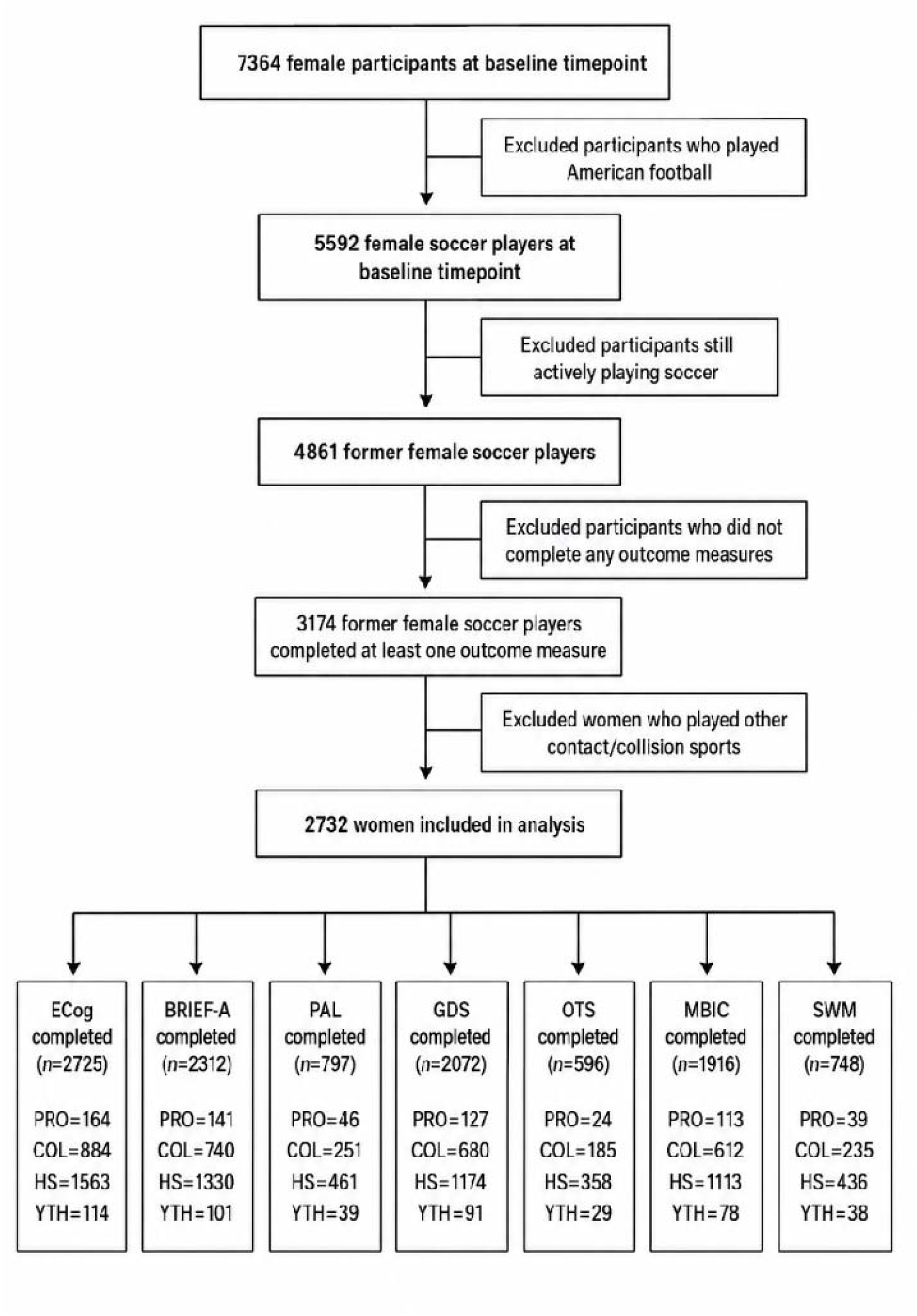
Sample derivation and task completion rates.

### 3.2 Relationships between RHI proxies and outcomes

#### 3.2.1 Level of Play

ANCOVA models demonstrated a significant omnibus effect for highest level of play on GDS-15 (F=3.79, p_adj_=0.030) and BRIEF BRI (F=3.06, p_adj_=0.041). **Table 2**. Post-hoc, Tukey-adjusted pairwise comparisons demonstrated that professional players scored significantly worse than high school players on both the GDS-15 (p=0.010) and BRIEF BRI (p=0.049), and also scored significantly worse than youth players on the BRIEF BRI (p=0.043) (**Figure 2).**

**Table 2.** Primary results: associations between RHI exposures and cognitive/neurobehavioral outcomes. P-values BH-corrected within domain × exposure.

| Outcome | Predictor | B (95% CI) or F-statistic | Adjusted P-value |
| --- | --- | --- | --- |
| PAL FAMS | Highest level of play | F(3, 792) = 0.95 | 0.623 |
|  | Total years of play | 0.00 (-0.05, 0.05) | 0.988 |
|  | Cumulative heading | -0.01 (-0.10, 0.08) | 0.908 |
|  | Concussion history | -0.07 (-0.11, -0.02) | 0.015 |
| PAL TEA | Highest level of play | F(3, 792) = 0.97 | 0.623 |
|  | Total years of play | 0.02 (-0.10, 0.14) | 0.988 |
|  | Cumulative heading | 0.05 (-0.17, 0.28) | 0.908 |
|  | Concussion history | 0.12 (0.01, 0.23) | 0.080 |
| OTSPSFC | Highest level of play | F(3, 591) = 0.79 | 0.623 |
|  | Total years of play | -0.00 (-0.04, 0.03) | 0.988 |
|  | Cumulative heading | 0.04 (-0.03, 0.11) | 0.661 |
|  | Concussion history | -0.01 (-0.05, 0.03) | 0.739 |
| SWM Between Errors | Highest level of play | F(3, 742) = 0.45 | 0.721 |
|  | Total years of play | 0.15 (0.02, 0.28) | 0.101 |
|  | Cumulative heading | 0.01 (-0.22, 0.25) | 0.908 |
|  | Concussion history | -0.03 (-0.14, 0.09) | 0.739 |
| SWM Strategy | Highest level of play | F(3, 742) = 2.21 | 0.430 |
|  | Total years of play | 0.02 (0.00, 0.04) | 0.101 |
|  | Cumulative heading | 0.02 (-0.01, 0.06) | 0.661 |
|  | Concussion history | -0.00 (-0.02, 0.01) | 0.739 |
| BRIEF MI | Highest level of play | F(3, 2324) = 1.08 | 0.355 |
|  | Total years of play | 0.02 (-0.09, 0.12) | 0.760 |
|  | Cumulative heading | 0.39 (0.19, 0.60) | <.001 |
|  | Concussion history | 0.24 (0.18, 0.30) | <.001 |
| ECog Mean | Highest level of play | F(3, 2717) = 1.30 | 0.355 |
|  | Total years of play | 0.00 (0.00, 0.01) | 0.083 |
|  | Cumulative heading | 0.02 (0.01, 0.03) | <.001 |
|  | Concussion history | 0.01 (0.01, 0.01) | <.001 |
| BRIEF BRI | Highest level of play | F(3, 2324) = 3.06 | 0.041 |
|  | Total years of play | 0.14 (0.05, 0.23) | 0.004 |
|  | Cumulative heading | 0.57 (0.38, 0.75) | <.001 |
|  | Concussion history | 0.22 (0.16, 0.28) | <.001 |
| GDS-15 | Highest level of play | F(3, 2075) = 3.79 | 0.030 |
|  | Total years of play | 0.02 (0.01, 0.04) | 0.002 |
|  | Cumulative heading | 0.06 (0.03, 0.09) | <.001 |
|  | Concussion history | 0.02 (0.01, 0.03) | <.001 |
| MBI-C Total | Highest level of play | F(3, 1224) = 1.09 | 0.354 |
|  | Total years of play | 0.14 (0.04, 0.24) | 0.005 |
|  | Cumulative heading | 0.40 (0.20, 0.60) | <.001 |
|  | Concussion history | 0.14 (0.09, 0.19) | <.001 |

Abbreviations: GDS-15 = Geriatric Depression Scale-15, ECog = Everyday Cognition Scale, PAL FAMS = Paired Associates Learning First Attempt Memory Score, PAL TEA = Paired Associates Learning Total Errors Adjusted, BRIEF-A = Behavior Rating Inventory of Executive Function – Adult, MI = Meta Cognition Index, BRI = Behavioral Regulation Index

**Figure 2:**
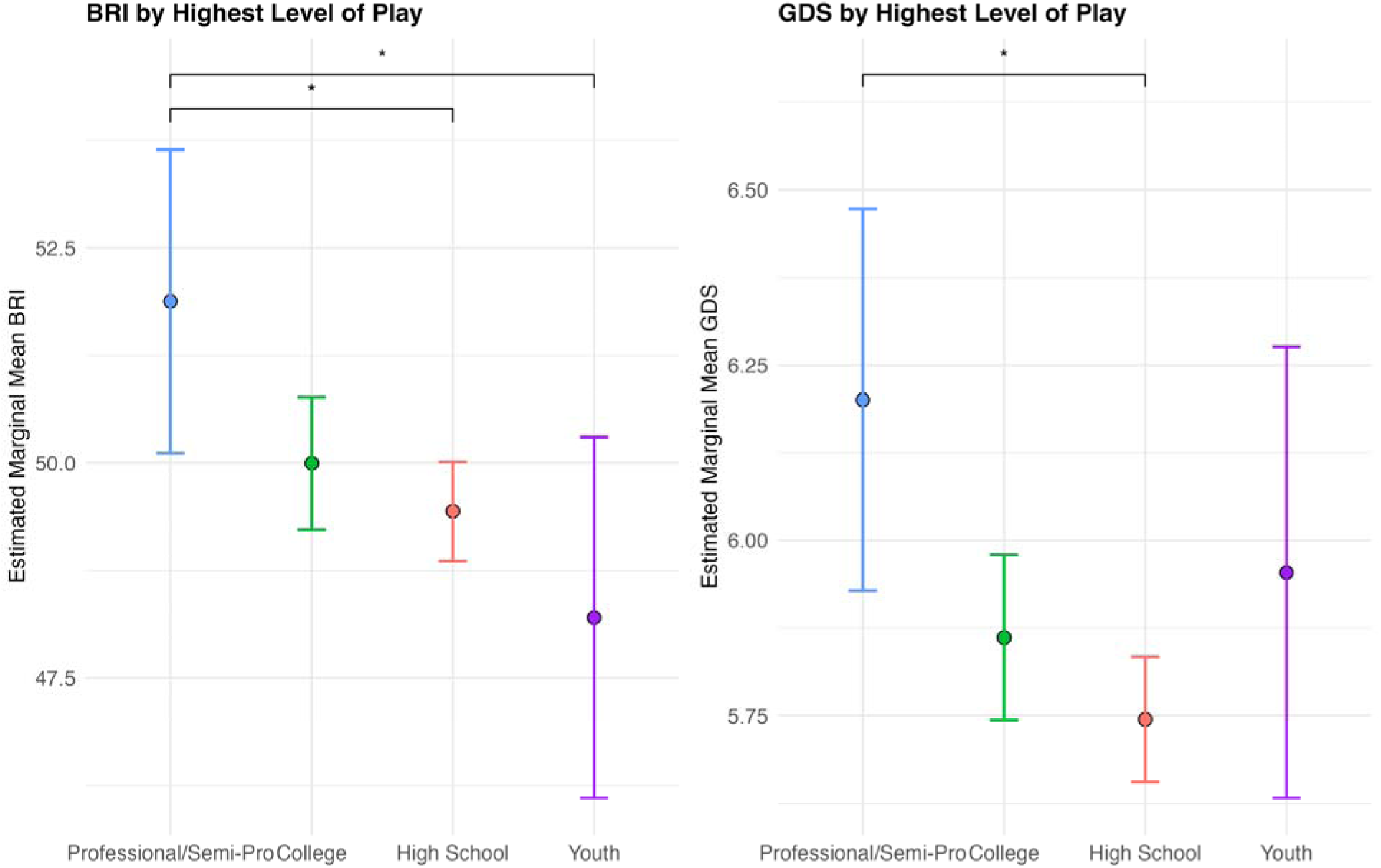
Estimated Marginal Mean Plots. Plots showing highest level of soccer play on the x-axis and estimated marginal means of outcome measures on the y-axis. An asterisk indicates a statistically significant difference from post-hoc, pairwise Tukey-adjusted comparisons

#### 3.2.3 Years of Play

Multivariable linear regression models demonstrated significant associations between more total years of soccer play and higher (worse) scores across several neurobehavioral and cognitive outcomes. Specifically, longer duration of play was associated with greater behavioral dysregulation on the BRIEF-A BRI (B = 0.14, 95% CI = [0.05, 0.23], p_adj_ = 0.004). More years of play was also significantly associated with worse scores on the GDS-15 (B = 0.02, 95% CI = [0.01, 0.04], p_adj_ = 0.002) and the MBI-C (B = 0.14, 95% CI = [0.04, 0.24], p_adj_ = 0.005).3.2.4 Estimated Cumulative Heading

Greater estimated cumulative heading was significantly associated with worse scores on the GDS-15 (B = 0.06, 95% CI = [0.03, 0.09, p_adj_ <0.001), the MBI-C (B = 0.40, 95% CI = [0.20, 0.60], p_adj_ <0.001) and the ECog (B = 0.02, 95% CI = [0.01, 0.03], p_adj_ <0.001). Furthermore, estimated cumulative heading was significantly associated with elevated scores on both global indices of the BRIEF MI (B= 0.39, 95% CI = [0.19, 0.60], padj<0.001), and the BRIEF BRI (B= 0.57, 95% CI = [0.38, 0.75], p_adj_ <0.001).

#### 3.2.5 Concussion History

Greater number of self-reported concussions was significantly associated with worse scores on the BRIEF MI (B = 0.24, 95% CI = [0.18, 0.30], p_adj_ <0.001), the BRIEF BRI (B = 0.22, 95% CI = [0.16, 0.28], p_adj_ <0.001), the ECog (B = 0.01, 95% CI = [0.01, 0.01], p_adj_ <0.001), the GDS-15 (B = 0.02, 95% CI = [0.01, 0.03], p_adj_ <0.001), and the MBI-C (B = 0.14, 95% CI = [0.09, 0.19], p_adj_ <0.001). Concussion history was also significantly associated with worse performance on the PAL FAMS (B = -0.07, 95% CI = [-0.11, -0.02], p_adj_ = 0.015). No significant associations were observed between concussion history and PAL TOTEA, SWM Strategy, or SWM Between Errors (all p_adj_ >0.05).

#### 3.3 Results of Sensitivity Analyses

##### 3.3.1: Analyses Excluding Pros

Excluding professional soccer players yielded smaller effect sizes for level of play, and associations with GDS-15 and BRIEF BRI lost significance altogether (Table 3). Associations with total years of play were diminished but remained significant for BRIEF BRI, GDS-15, and MBI-C, while effect sizes and significance patterns for estimated cumulative heading were largely unchanged. The magnitude of the effect sizes for concussion history increased upon excluding the professional players, while retaining statistical significance; for example, B increased from 0.24 to 0.60 for the BRIEF MI and 0.14 to 0.43 for the MBI-C.

**Table 3.**
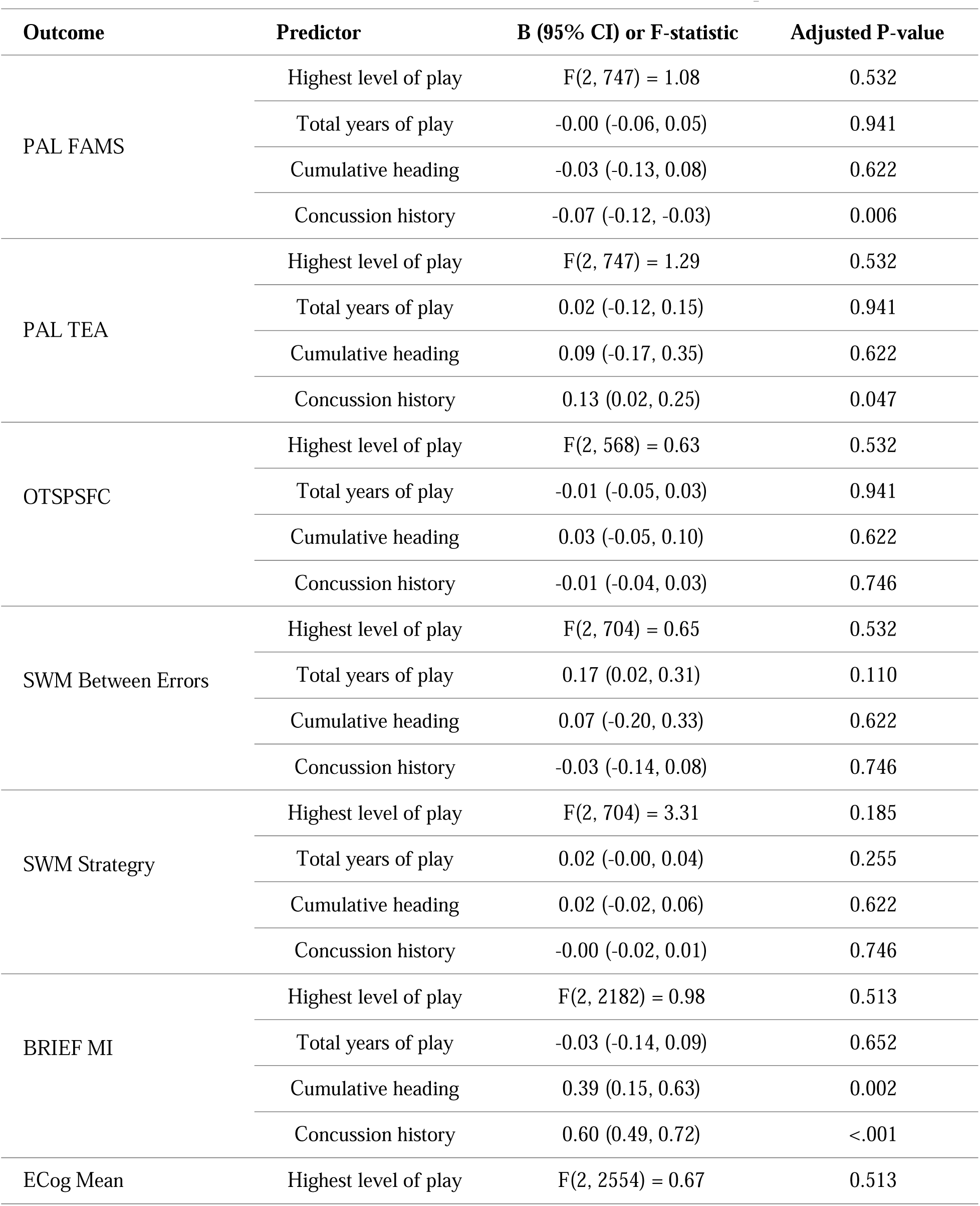
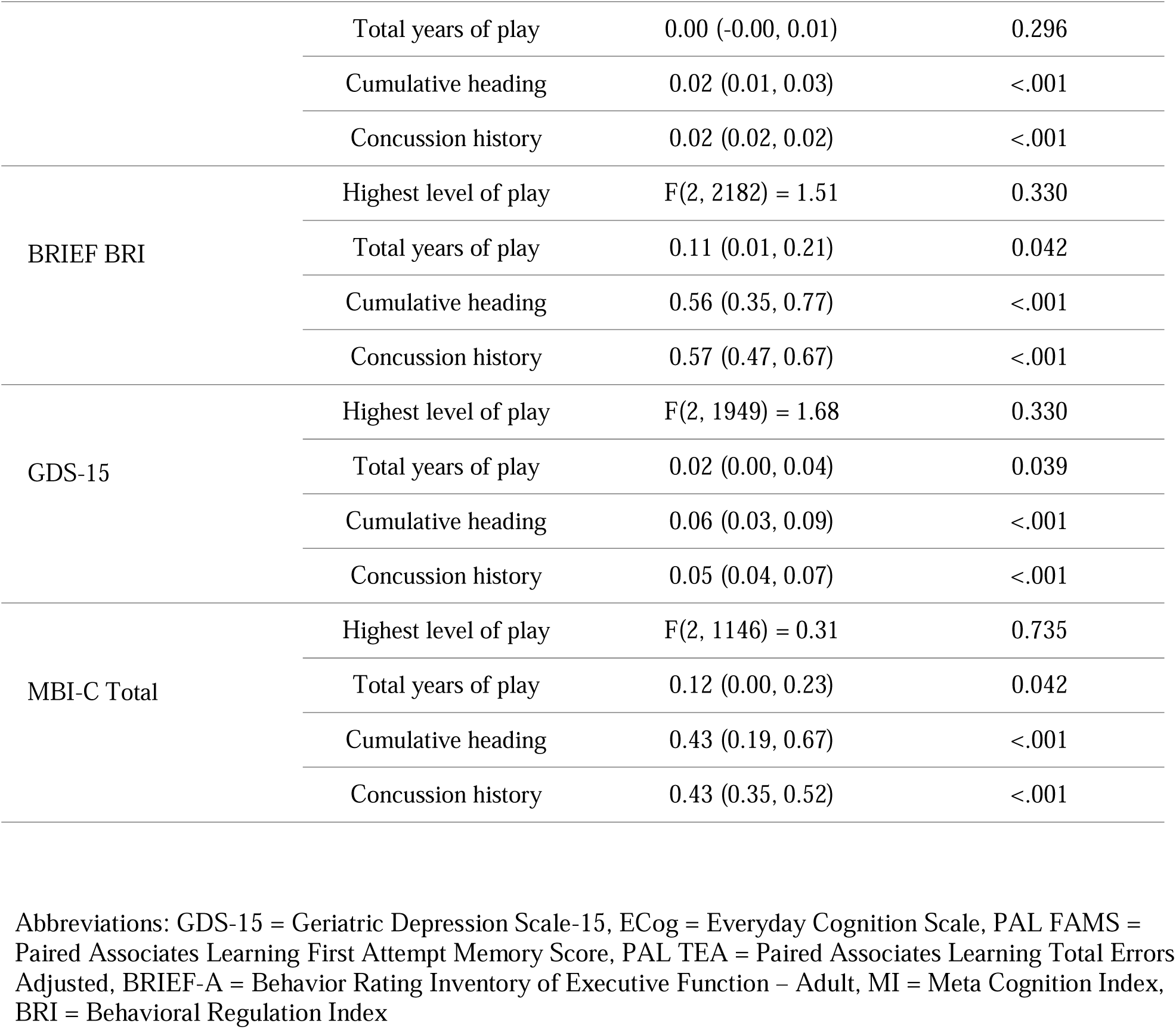
Sensitivity analysis excluding professional/semi-pro players. P-values BH-corrected within domain × exposure.

##### 3.3.2: Analyses of BRIEF Subscales

Exploratory multivariable linear regression models examining associations between the RHI exposure metrics and BRIEF scales demonstrated significant associations between concussion history and estimated cumulative heading with each outcome measure. (Table 4). BRIEF Inhibit and Emotional Control were significantly associated with all four predictors.

**Table 4.** Exploratory BRIEF subscale results. Uncorrected p-values; not adjusted for multiple comparisons.

| Outcome | Predictor | B (95% CI) or F-statistic | P-value |
| --- | --- | --- | --- |
| BRIEF Organization of Materials | Highest level of play | F(3, 2324) = 0.52 | 0.671 |
|  | Total years of play | -0.01 (-0.11, 0.09) | 0.786 |
|  | Cumulative heading | 0.24 (0.04, 0.44) | 0.021 |
|  | Concussion history | 0.18 (0.12, 0.24) | <.001 |
| BRIEF Task Monitor | Highest level of play | F(3, 2324) = 2.19 | 0.088 |
|  | Total years of play | -0.01 (-0.11, 0.09) | 0.822 |
|  | Cumulative heading | 0.31 (0.12, 0.51) | 0.002 |
|  | Concussion history | 0.21 (0.15, 0.27) | <.001 |
| BRIEF Plan/Organize | Highest level of play | F(3, 2323) = 1.55 | 0.200 |
|  | Total years of play | 0.00 (-0.09, 0.10) | 0.922 |
|  | Cumulative heading | 0.32 (0.12, 0.51) | 0.001 |
|  | Concussion history | 0.22 (0.16, 0.28) | <.001 |
| BRIEF Working Memory | Highest level of play | F(3, 2324) = 0.86 | 0.463 |
|  | Total years of play | 0.08 (-0.03, 0.19) | 0.146 |
|  | Cumulative heading | 0.57 (0.36, 0.79) | <.001 |
|  | Concussion history | 0.24 (0.17, 0.30) | <.001 |
| BRIEF Initiate | Highest level of play | F(3, 2324) = 1.59 | 0.190 |
|  | Total years of play | 0.02 (-0.08, 0.12) | 0.732 |
|  | Cumulative heading | 0.29 (0.09, 0.49) | 0.005 |
|  | Concussion history | 0.19 (0.13, 0.25) | <.001 |
| BRIEF Self-Monitor | Highest level of play | F(3, 2323) = 1.50 | 0.212 |
|  | Total years of play | 0.06 (-0.02, 0.14) | 0.133 |
|  | Cumulative heading | 0.34 (0.18, 0.50) | <.001 |
|  | Concussion history | 0.18 (0.13, 0.22) | <.001 |
| BRIEF Shift | Highest level of play | F(3, 2324) = 1.34 | 0.260 |
|  | Total years of play | 0.07 (-0.02, 0.16) | 0.137 |
|  | Cumulative heading | 0.44 (0.26, 0.63) | <.001 |
|  | Concussion history | 0.17 (0.12, 0.23) | <.001 |
| BRIEF Inhibit | Highest level of play | F(3, 2324) = 2.70 | 0.044 |
|  | Total years of play | 0.14 (0.05, 0.22) | 0.001 |
|  | Cumulative heading | 0.51 (0.34, 0.68) | <.001 |
|  | Concussion history | 0.20 (0.15, 0.25) | <.001 |
| BRIEF Emotional Control | Highest level of play | F(3, 2324) = 3.07 | 0.027 |
|  | Total years of play | 0.14 (0.05, 0.24) | 0.002 |
|  | Cumulative heading | 0.49 (0.31, 0.68) | <.001 |
|  | Concussion history | 0.17 (0.11, 0.23) | <.001 |

BRIEF-A = Behavior Rating Inventory of Executive Function

### 4.0 DISCUSSION

This study evaluated associations between proxies of RHI exposure in soccer and measures of cognition, subjective cognitive complaints, behavioral dysregulation and depressive symptoms among a large cohort of female former soccer players, age 40 and older, spanning amateurs through professionals. We found small but statistically significant associations between greater estimated cumulative heading exposure, higher level of play, total years of play, and concussion history, and worse subjective cognitive complaints, self-reported behavioral dysregulation, and depressive symptom severity. No significant associations were found between RHI proxies and objective computerized cognitive test performance, apart from PAL FAMS and concussion history. To our knowledge, this represents the largest investigation of clinical symptoms and RHI exposure among middle-aged female former soccer players across all levels of play.

We found that both greater estimated cumulative heading exposure and increased concussions were associated with worse scores on most of the self-report measures of subjective cognitive complaints, depressive symptoms, and behavioral dysregulation, although overall effect sizes were small. Previous studies demonstrated adverse neurocognitive effects of heading the ball in soccer.^51^ For example, Bruno et al found that, among male former professional soccer players, higher estimated total career headers were associated with lower scores on Test Your Memory, a brief cognitive screening instrument.^52^ Prior studies have also shown high prevalence of subjective cognitive complaints and decline among former athletes, including former football players^53^ and soccer players.^32^ Furthermore, while numerous studies have observed associations between increased heading frequency and both acute and later-life impairments, the neurobiological mechanisms underpinning these impairments remain unclear. A recent study identified the orbitofrontal gray-white matter interface as a locus of RHI-related injury in soccer, associated with poorer verbal learning.^54^ Similarly, a systematic review of sport-related concussion and neurocognitive outcomes among former athletes, including soccer players, revealed elevated self-report cognitive impairments.^55^ Prospective, large-scale investigation into pathological changes associated with heading and concussion, and the association of these changes with clinical symptoms is warranted.

We found some significant associations between higher level of soccer play as well as longer duration of play, and elevations on self-report measures of behavioral dysregulation and depressive symptoms. Neurobehavioral dysregulation (e.g., explosivity, impulsivity, emotional lability) is a core clinical feature of traumatic encephalopathy syndrome (TES),^56^ the proposed clinical syndrome associated with CTE pathology. Although depression is not a core clinical feature, it is one of several supportive features for determining levels of certainty for CTE pathology in the TES diagnostic framework. It is important to note, however, that TES features have not all been systematically verified against autopsy-confirmed CTE neuropathology. In a clinicopathological validation study of 336 donors, cognitive symptoms were significantly associated with CTE pathology, while mood and behavioral symptoms were not.^57^ Neurobehavioral dysregulation, in isolation, therefore does not reliably predict underlying CTE neuropathologic change. Neurobehavioral dysregulation has been examined in male football players,^58–62^ but has not yet been characterized in women at risk of CTE. This is crucial not only in terms of sex-based differential vulnerability to head impacts,^63,64^ but also in terms of discrepancies in how women experience and self-report symptoms. For example, Schaffert and colleagues found that emotional symptoms were a stronger predictor of self-reported cognitive symptoms than objective impairment among older retired female collegiate athletes.^65^ In this vein, the relationship between depressive symptoms and subjective complaints warrants further consideration. Depression in middle age could be a major factor accounting for cognitive and behavioral complaints among this cohort. This underscores the need for longitudinal monitoring of disease trajectories.

We found few associations between RHI proxies and the computerized CANTAB tests used in this study as measures of objective cognitive impairment. This could be due to multiple factors, namely, that our cohort was young, mostly cognitively unimpaired, and exhibited no real objective impairments. Symptoms of any possible neurodegenerative conditions may not manifest until later in life. Additionally, there were relatively low completion rates for the CANTAB measures. A recent study examining feasibility, acceptability, and construct validity of CANTAB PAL in BHR found that individuals with subjective memory concerns were less likely to complete the assessment.^42^ Disproportionate completion of the measures could have biased our CANTAB findings toward the null.

Null findings and small effect sizes in our study could also stem from the ways in which RHI exposure was operationalized in the study. Unlike in American football, where helmet accelerometer data has validated metrics like the Cumulative Head Impact Index, ^2,66^ soccer lacks such measures. Though there has been some usage of headbands and other sensors in soccer,^67^ there are no existing datasets that could provide adequate data by season, for specific positions and levels of play. Additionally, total years of play is a crude metric for ascertaining exposure patterns in soccer, since there is substantial variability in terms of competitive structures and individual players’ total seasons played per year. RHI exposure in soccer is also heterogeneous. For example, heading frequency varies substantially across position of play, play style, and physical attributes (e.g., height), among other factors.^68,69^ Consequently, the symptom burden observed in our cohort may be driven by a subset of athletes with high cumulative exposure, rather than signifying uniform risk. This heterogeneity could help explain why estimated cumulative heading was a sensitive predictor in our models, as it may capture individual behavior more effectively than other RHI proxies. Ultimately, this demonstrates that RHI exposure metrics used in football may not effectively quantify the spectrum of exposures in soccer.

### Limitations

Our study has several limitations. Multiple selection biases limit the generalizability of our findings, including lack of ethnocultural and educational diversity in the sample, requirement for an internet-connected device and digital literacy, and underrepresentation of players whose highest level of play was youth. Recreational soccer play was excluded from the analysis, therefore eliminating an additional dimension of exposure. College soccer players only included varsity and not club or intramural players. Importantly, this study failed to include an appropriate, non-RHI exposed control group for comparison across outcomes. High prevalence of depressive symptoms and subjective cognitive concerns within this cohort could reflect selection bias for online Alzheimer’s disease and related dementias research registries.

Only three CANTAB measures were administered in the HITSS battery, which assessed episodic memory and visual learning, spatial planning, and spatial working memory. There are limitations to unsupervised, computerized cognitive testing, including lack of free response and lack of supervision. Unmeasured, confounding factors such as family history of neurodegenerative diseases, physical and psychiatric comorbidities, and sociocultural and structural determinants of health, all of which could influence cognitive and neurobehavioral outcomes, but that weren’t considered in this analysis. HITSS and BHR questionnaires were self-report and therefore may be subject to error and recall bias. Estimated cumulative heading was operationalized as an ordinal measure, and future research is needed to determine whether heading frequency varies significantly across different levels of soccer play. Lastly, missing data, especially relatively low completion rates for the CANTAB measures could have limited power to detect significant effects.

It is important to consider the evolution of soccer conduct over the past several decades, including implementation of mandates to prohibit heading for players aged 10 and younger in the United States. Age of first exposure (AFE) has been examined as an RHI exposure metric primarily in American football. Few studies have examined AFE in soccer. One prior study included a small number of younger, active players and found no associations.^70^ Interestingly, another study of adult amateur soccer players found that earlier AFE (≤10 years) was associated with better performance on working memory and verbal learning tasks in young adulthood, with no adverse associations observed.^71^ However, AFE may be a flawed metric in soccer since first exposure to the sport may not coincide with AFE to RHI. Subsequent studies should collect information specifically about AFE to heading.

Another important consideration is the transition from leather soccer balls, which were prone to weight increase from water absorption,^72^ to hydrophobic synthetic balls beginning in the 1960s, but only becoming the dominant standard at elite levels in the 1990s. Furthermore, Title IX of the Education Amendments of 1972 (Title IX) mandated equal opportunities for men and women in sports. Since then, the percentage of female athletes has significantly exponentially increased^73,74^ at the high school, college, and professional levels. At the college level, it is estimated that over 4 million women have played college soccer since the passing of Title IX, according to the National Collegiate Athletic Association, with close to 30,000 women playing soccer annually at the Division I, II, and III levels.^13^ As the first wave of Title IX athletes age, the number of women at risk for aging-related neurologic conditions is expected to increase. Because of the slow growth of sports opportunities after the implementation of Title IX, studies of male and female soccer players may not be comparable due to exposure differences not captured by measures like years of play. The older women in this study lacked the same professional opportunities provided to men, both in length and intensity of their RHI exposure. In addition, the younger subjects in this study were more likely to play when soccer participation was year-round rather than only for one or two seasons a year, which may mask the cumulative effects of RHI and aging seen in other HITSS publications. Therefore, understanding later-life consequences of soccer play among women is of critical importance.

### Conclusions

In conclusion, we found that, within a cohort of female former soccer players, there were significant associations between higher RHI exposure and self-report cognitive, neurobehavioral, and depression-related outcomes. There was only one association between an RHI exposure metric and objective computerized neuropsychological test. Future analyses should examine more granular RHI exposure metrics, as well as longitudinal change in the neurocognitive and behavioral outcome measures, and whether these changes are associated with fluid and neuroimaging biomarkers of underlying pathophysiological change and neurodegenerative disease.

## Data Availability

Data will be available upon request via the Federal Interagency Traumatic Brain Injury Research (FITBIR) database

https://fitbir.nih.gov/

## AUTHORS’ CONTRIBUTIONS

SCM and AA contributed equally and have shared first authorship as the primary authors who were involved in data analysis and drafting the manuscript

Funding Acquisition: RAS, MLA, CJN, ACM

Project administration: SCM, KJG, WCSF, MR, GS, KHZ, ACM, MDM, RLN

Writing – original draft: SCM, AA, RAS

Curation of data: JNP, BMM, GS, BNP

Writing – review & editing: FTZ, KJG, YT, WCSF, GS, JNP, BMM, BNP, JNP, BMM, KHZ, DIK, CJN, ACM, TDS, RSM, MDM, JW, JM, MWW, RLN, MLA, RAS

Formal Analysis: YT, FTZ, AA, SCM

Supervision: JNP, ACM, RAS, MLA

## CONSENT TO PARTICIPATE

All participants were required to sign an Informed Consent Form as approved by the IRB prior to enrolling in the study.

## FIGURE TITLES AND LEGENDS

Figure 1: HITSS soccer cohort derivation and task completion

Figure 2: Cognitive and Neurobehavioral Outcomes by Highest Level of Soccer Play

Figure 2 Caption: Plots showing highest level of soccer play on the x-axis and estimated marginal means of outcome measures on the y-axis.

## References

1. Alosco ML, Barr WB, Banks SJ, et al. Neuropsychological test performance of former American football players. Alzheimers Res Ther 2023; 15: 1.

2. Montenigro PH, Alosco ML, Martin BM, et al. Cumulative Head Impact Exposure Predicts Later-Life Depression, Apathy, Executive Dysfunction, and Cognitive Impairment in Former High School and College Football Players. J Neurotrauma 2017; 34: 328–340.

3. Phelps A, Alosco ML, Baucom Z, et al. Association of Playing College American Football With Long-term Health Outcomes and Mortality. JAMA Netw Open 2022; 5: e228775.

4. Schaffert J, LoBue C, Fields L, et al. Neuropsychological functioning in ageing retired NFL players: a critical review. Int Rev Psychiatry 2020; 32: 71–88.

5. McKee AC, Mez J, Abdolmohammadi B, et al. Neuropathologic and Clinical Findings in Young Contact Sport Athletes Exposed to Repetitive Head Impacts. JAMA Neurol 2023; 80: 1037–1050.

6. McKee AC, Stein TD, Huber BR, et al. Chronic traumatic encephalopathy (CTE): criteria for neuropathological diagnosis and relationship to repetitive head impacts. Acta Neuropathol (Berl) 2023; 145: 371–394.

7. van Amerongen S, Kamps S, Kaijser KKM, et al. Severe CTE and TDP-43 pathology in a former professional soccer player with dementia: a clinicopathological case report and review of the literature. Acta Neuropathol Commun 2023; 11: 77.

8. Ling H, Morris HR, Neal JW, et al. Mixed pathologies including chronic traumatic encephalopathy account for dementia in retired association football (soccer) players. Acta Neuropathol (Berl) 2017; 133: 337–352.

9. Grinberg LT, Anghinah R, Nascimento CF, et al. Chronic Traumatic Encephalopathy Presenting as Alzheimer’s Disease in a Retired Soccer Player. J Alzheimer’s Dis 2016; 54: 169–174.

10. Ueda P, Pasternak B, Lim C-E, et al. Neurodegenerative disease among male elite football (soccer) players in Sweden: a cohort study. Lancet Public Health 2023; 8: e256–e265.

11. DeMessie B, Stewart WF, Lipton RB, et al. Soccer Heading Exposure–Dependent Microstructural Injury at Depths of Sulci in Adult Amateur Players. Neurology 2025; 105: e214034.

12. Yao Z-F, Sligte IG, Moreau D, et al. The brains of elite soccer players are subject to experience-dependent alterations in white matter connectivity. Cortex J Devoted Study Nerv Syst Behav 2020; 132: 79–91.

13. Koerte IK, Lin AP, Muehlmann M, et al. Altered Neurochemistry in Former Professional Soccer Players without a History of Concussion. J Neurotrauma 2015; 32: 1287–1293.

14. Levitch CF, Zimmerman ME, Lubin N, et al. Recent and Long-Term Soccer Heading Exposure Is Differentially Associated With Neuropsychological Function in Amateur Players. J Int Neuropsychol Soc JINS 2018; 24: 147–155.

15. Zhang MR, Red SD, Lin AH, et al. Evidence of Cognitive Dysfunction after Soccer Playing with Ball Heading Using a Novel Tablet-Based Approach. PLOS ONE 2013; 8: e57364.

16. Ashton J, Coyles G, Malone JJ, et al. Immediate effects of an acute bout of repeated soccer heading on cognitive performance. Sci Med Footb 2021; 5: 181–187.

17. Hoppen MI, Königs M, Teunissen CE, et al. Amateur Soccer Heading and Acute Elevations in Blood-Based p-Tau217 and S100B. JAMA Neurol. Epub ahead of print 18 May 2026. DOI: 10.1001/jamaneurol.2026.1224.

18. McLean C, Lavender AP, Pereira E, et al. The Acute Effects of Ball Pressure on Anticipation Timing Following a Series of Purposeful Headers in Adult Football (Soccer) Players. Sports Basel Switz 2024; 12: 102.

19. Zetterberg H, Jonsson M, Rasulzada A, et al. No neurochemical evidence for brain injury caused by heading in soccer. Br J Sports Med 2007; 41: 574–577.

20. Levitch CF, Zimmerman ME, Lubin N, et al. Recent and Long-Term Soccer Heading Exposure Is Differentially Associated With Neuropsychological Function in Amateur Players. J Int Neuropsychol Soc JINS 2018; 24: 147–155.

21. DeMessie B, Charney MF, Fleysher R, et al. Soccer heading and white matter microstructural changes: a two-year longitudinal cohort study. Brain Imaging Behav 2026; 20: 52.

22. Lipton ML, Kim N, Zimmerman ME, et al. Soccer Heading Is Associated with White Matter Microstructural and Cognitive Abnormalities. Radiology 2013; 268: 850–857.

23. Russell ER, Mackay DF, Stewart K, et al. Association of Field Position and Career Length With Risk of Neurodegenerative Disease in Male Former Professional Soccer Players. JAMA Neurol 2021; 78: 1057–1063.

24. Mackay DF, Russell ER, Stewart K, et al. Neurodegenerative Disease Mortality among Former Professional Soccer Players. N Engl J Med 2019; 381: 1801–1808.

25. Fahr J, Kraff O, Deuschl C, et al. Concussion in Female Athletes of Contact Sports: A Scoping Review. Orthop J Sports Med 2024; 12: 23259671241276447.

26. Powell JW, Barber-Foss KD. Traumatic Brain Injury in High School Athletes. JAMA 1999; 282: 958–963.

27. Covassin T, Elbin RJ, Bleecker A, et al. Are there differences in neurocognitive function and symptoms between male and female soccer players after concussions? Am J Sports Med 2013; 41: 2890–2895.

28. Rubin TG, Catenaccio E, Fleysher R, et al. MRI-defined White Matter Microstructural Alteration Associated with Soccer Heading Is More Extensive in Women than Men. Radiology 2018; 289: 478–486.

29. Caccese JB, Buckley TA, Tierney RT, et al. Head and neck size and neck strength predict linear and rotational acceleration during purposeful soccer heading. Sports Biomech 2018; 17: 462–476.

30. Haynes N, Goodwin T. Literature Review of Sex Differences in mTBI. Mil Med 2023; 188: e978–e984.

31. Haase FK, Prien A, Douw L, et al. Cortical thickness and neurocognitive performance in former high-level female soccer and non-contact sport athletes. Scand J Med Sci Sports 2023; 33: 921–930.

32. Prien A, Feddermann-Demont N, Verhagen E, et al. Neurocognitive performance and mental health of retired female football players compared to non-contact sport athletes. BMJ Open Sport Exerc Med 2020; 6: e000952.

33. Kern J, Gulde P, Hermsdörfer J. A prospective investigation of the effects of soccer heading on cognitive and sensorimotor performances in semi-professional female players. Front Hum Neurosci 2024; 18: 1345868.

34. Prien A, Besuden C, Junge A, et al. Cognitive Ageing in Top-Level Female Soccer Players Compared to a Normative Sample from the General Population: A Cross-sectional Study. J Int Neuropsychol Soc JINS 2020; 26: 645–653.

35. Weiner MW, Nosheny R, Camacho M, et al. The Brain Health Registry: An internet-based platform for recruitment, assessment, and longitudinal monitoring of participants for neuroscience studies. Alzheimers Dement J Alzheimers Assoc 2018; 14: 1063–1076.

36. Weiner MW, Aaronson A, Eichenbaum J, et al. Brain health registry updates: An online longitudinal neuroscience platform. Alzheimers Dement J Alzheimers Assoc 2023; 19: 4935–4951.

37. Mez J, Solomon TM, Daneshvar DH, et al. Assessing clinicopathological correlation in chronic traumatic encephalopathy: rationale and methods for the UNITE study. Alzheimers Res Ther 2015; 7: 62.

38. Alosco ML, Mian AZ, Buch K, et al. Structural MRI profiles and tau correlates of atrophy in autopsy-confirmed CTE. Alzheimers Res Ther 2021; 13: 193.

39. Bruce HJ, Tripodis Y, McClean M, et al. American Football Play and Parkinson Disease Among Men. JAMA Netw Open 2023; 6: e2328644.

40. Roth RM, Isquith PK, Gioia GA. Behavior Rating Inventory of Executive Function – Adult Version (BRIEF-A). Lutz, FL, 2005.

41. Farias ST, Mungas D, Reed BR, et al. The measurement of everyday cognition (ECog): scale development and psychometric properties. Neuropsychology 2008; 22: 531–544.

42. Ashford MT, Aaronson A, Kwang W, et al. Unsupervised Online Paired Associates Learning Task from the Cambridge Neuropsychological Test Automated Battery (CANTAB®) in the Brain Health Registry. J Prev Alzheimers Dis 2024; 11: 514–524.

43. Pettigrew C, Soldan A, Brichko R, et al. Computerized paired associate learning performance and imaging biomarkers in older adults without dementia. Brain Imaging Behav 2022; 16: 921–929.

44. Barnett JH, Blackwell AD, Sahakian BJ, et al. The Paired Associates Learning (PAL) Test: 30 Years of CANTAB Translational Neuroscience from Laboratory to Bedside in Dementia Research. Curr Top Behav Neurosci 2016; 28: 449–474.

45. Digital cognitive assessments. Cambridge Cognition, https://cambridgecognition.com/digital-cognitive-assessments/ (accessed 16 January 2026).

46. Gioia GA, Isquith PK, Guy SC, et al. TEST REVIEW Behavior Rating Inventory of Executive Function. Child Neuropsychol. Epub ahead of print 1 September 2000. DOI: 10.1076/chin.6.3.235.3152.

47. Sheikh JI, Yesavage JA. Geriatric Depression Scale (GDS): Recent evidence and development of a shorter version. Clin Gerontol J Aging Ment Health 1986; 5: 165–173.

48. Guerin JM, Copersino ML, Schretlen DJ. Clinical utility of the 15-item geriatric depression scale (GDS-15) for use with young and middle-aged adults. J Affect Disord 2018; 241: 59–62.

49. Ismail Z, Agüera-Ortiz L, Brodaty H, et al. The Mild Behavioral Impairment Checklist (MBI-C): A Rating Scale for Neuropsychiatric Symptoms in Pre-Dementia Populations. J Alzheimers Dis JAD 2017; 56: 929–938.

50. Mallo SC, Ismail Z, Pereiro AX, et al. Assessing mild behavioral impairment with the mild behavioral impairment checklist in people with subjective cognitive decline. Int Psychogeriatr 2019; 31: 231–239.

51. Spiotta AM, Bartsch AJ, Benzel EC. Heading in soccer: dangerous play? Neurosurgery 2012; 70: 1–11; discussion 11.

52. Bruno D, Rutherford A. Cognitive ability in former professional football (soccer) players is associated with estimated heading frequency. J Neuropsychol 2022; 16: 434–443.

53. Brett BL, Kerr ZY, Chandran A, et al. A dominance analysis of subjective cognitive complaint comorbidities in former professional football players with and without mild cognitive impairment. J Int Neuropsychol Soc JINS 2023; 29: 582–593.

54. Song JY, Fleysher R, Ye K, et al. Orbitofrontal Gray-White Interface Injury and the Association of Soccer Heading With Verbal Learning. JAMA Netw Open 2025; 8: e2532461.

55. Cunningham J, Broglio SP, O’Grady M, et al. History of Sport-Related Concussion and Long-Term Clinical Cognitive Health Outcomes in Retired Athletes: A Systematic Review. J Athl Train 2020; 55: 132–158.

56. Katz DI, Bernick C, Dodick DW, et al. National Institute of Neurological Disorders and Stroke Consensus Diagnostic Criteria for Traumatic Encephalopathy Syndrome. Neurology 2021; 96: 848–863.

57. Mez J, Alosco ML, Daneshvar DH, et al. Validity of the 2014 traumatic encephalopathy syndrome criteria for CTE pathology. Alzheimers Dement 2021; 17: 1709–1724.

58. Pulukuri SV, Fagle TR, Trujillo-Rodriguez D, et al. Characterizing Neurobehavioral Dysregulation Among Former American Football Players: Findings From the DIAGNOSE CTE Research Project. J Neuropsychiatry Clin Neurosci 2025; 37: 38–46.

59. van Amerongen S, Pulukuri SV, Tuz-Zahra F, et al. Inflammatory biomarkers for neurobehavioral dysregulation in former American football players: findings from the DIAGNOSE CTE Research Project. J Neuroinflammation 2024; 21: 46.

60. van Amerongen S, Peskind ER, Tripodis Y, et al. Catecholamine Dysregulation in Former American Football Players. Neurology 2025; 104: e213584.

61. Wiegand TLT, Pankatz L, Arciniega H, et al. Diffusion Alterations at the Gray Matter/White Matter Boundary in Traumatic Encephalopathy Syndrome. J Neurotrauma. Epub ahead of print 10 November 2025. DOI: 10.1177/08977151251393966.

62. Jansson D, Shofer J, Colasurdo E, et al. Alterations in CSF Amyloid-β and Tau Biomarkers in Former College and Professional American Football Players: Findings from the DIAGNOSE CTE Research Project. J Neurotrauma 2026; 08977151251390520.

63. Gupte R, Brooks W, Vukas R, et al. Sex Differences in Traumatic Brain Injury: What We Know and What We Should Know. J Neurotrauma 2019; 36: 3063–3091.

64. Sandhu MRS, Schonwald A, Boyko M, et al. The association between female sex and depression following traumatic brain injury: A systematic review and meta-analysis. Neurosci Biobehav Rev 2025; 168: 105952.

65. Schaffert J, Sanders GD, Wilmoth K, et al. Repetitive Head-Impact Exposure and Cognitive and Emotional Symptoms in Older Retired Female Collegiate Athletes. J Head Trauma Rehabil 2025; 40: 440–447.

66. Daneshvar DH, Nair ES, Baucom ZH, et al. Leveraging football accelerometer data to quantify associations between repetitive head impacts and chronic traumatic encephalopathy in males. Nat Commun 2023; 14: 3470.

67. Deju B, Afzal H, Basnyat S, et al. Accelerometer-based head impact detection in soccer - Where are we? Health Sci Rev 2024; 10: 100141.

68. Alexander J, Gillett M, Patel S, et al. Quantification of heading in adult football: a systematic review and evidence synthesis. Epub ahead of print 1 December 2025. DOI: 10.1136/bjsports-2024-109462.

69. Brownlow M, Townsend DC, Varley I, et al. Heading in football matches: descriptive analysis of heading events from 687 matches across age groups in English Academy and First Team men’s professional football. Br J Sports Med 2025; 59: e110804.

70. Caccese JB, Santos FV, Yamaguchi F, et al. Age of First Exposure to Soccer Heading and Sensory Reweighting for Upright Stance. Int J Sports Med 2020; 41: 616–627.

71. Charney MF, Ye KQ, Fleysher R, et al. Age of first exposure to soccer heading: Associations with cognitive, clinical, and imaging outcomes in the Einstein Soccer Study. Front Neurol 2023; 14: 1042707.

72. Phillips I, Harland A, Mitchell S, et al. Investigating static water uptake characteristics and mechanisms for leather and synthetic association footballs. Sports Eng 2025; 28: 40.

73. Gregg EA, Gregg VH. Women in Sport: Historical Perspectives. Clin Sports Med 2017; 36: 603–610.

74. O’Connor MI. Equity360: Gender, Race, and Ethnicity—Title IX Turns 50: Women Athletes Are Still Fighting Against Gender Disparities in Sports. Clin Orthop 2022; 480: 29–32.

